# A response playbook for early detection and population surveillance of new SARS-CoV-2 variants in a regional public health laboratory

**DOI:** 10.1101/2023.09.01.23293209

**Authors:** Hannah J. Barbian, Alyse Kittner, Richard Teran, Sofiya Bobrovska, Xueting Qiu, Kayla English, Stefan J. Green, Isaac Ghinai, Massimo Pacilli, Mary K. Hayden

## Abstract

**Background:** Timely genomic surveillance is required to inform public health responses to new SARS-CoV-2 variants. However, the processes involved in local genomic surveillance introduce inherent time constraints. The Regional Innovative Public Health Laboratory in Chicago developed and employed a genomic surveillance response playbook for the early detection and surveillance of emerging SARS-CoV-2 variants.

**Methods:** The playbook outlines modifications to sampling strategies, laboratory workflows, and communication processes based on the emerging variant’s predicted viral characteristics, observed public health impact in other jurisdictions and local community risk level. The playbook outlines procedures for implementing and reporting enhanced and accelerated genomic surveillance, including supplementing whole genome sequencing (WGS) with variant screening by quantitative PCR (qPCR).

**Results:** The ability of the playbook to improve the response to an emerging variant was tested for SARS-CoV-2 Omicron BA.1. Increased submission of clinical remnant samples from local hospital laboratories enabled detection of a new variant at 1% prevalence with 95% confidence rather than 2% at baseline. Genotyping qPCR concurred with WGS lineage assignments in 99.9% of 1541 samples with results by both methods, and was more sensitive, providing lineage results in 90.4% of 1833 samples rather than 85.1% for WGS, while reducing the time to lineage result from 27 to 7 days.

**Conclusions:** The genomic surveillance response playbook provides a structured, stepwise, and data-driven approach to responding to emerging SARS-CoV-2 variants. These pre-defined processes can streamline workflows and expedite the detection and public health response to emerging variants. Based on the processes piloted during the Omicron BA.1 response, this method has been applied to subsequent Omicron subvariants and can be readily applied to future SARS-CoV-2 emerging variants and other public health surveillance activities.

## Background

Like most of the United States, Chicago has experienced continued and successive waves of COVID-19 transmission driven in part by the emergence of new SARS-CoV-2 lineages with increased transmissibility.^1^ Timely and accurate estimates of local variant prevalence is required for the public health response to emerging SARS-CoV-2 variants of concern (VOCs)^2^ and has informed public health guidance on the use of monoclonal antibody treatment and non-pharmaceutical interventions such as indoor mask requirements.^3^ Additionally, sequencing SARS-CoV-2 genomes provides insight on transmission and biological and epidemic features of the VOCs that can inform public health recommendations and enhances traditional public health outbreak response by strengthening inferred transmission links.^4-5^

Whole-genome sequencing (WGS) is the gold standard for monitoring SARS-CoV-2 lineages^6^, however, it is labor intensive, expensive, has intrinsic limits on processing time, and can be logistically challenging to scale up.^7^ Other methods, such as genotyping PCR, are quicker, less expensive and can be performed at non-specialized laboratories.^8,9^ Rapid high-level genotyping data can provide more timely variant prevalence estimates prior to WGS data availability, as was observed by chance in variants B.1.1.7 and BA.1, which were more rapidly tracked via S-gene target failure with particular diagnostic assays due to a particular spike deletion.^10, 11, 12^

Genomic surveillance programs often require collaborations with existing public health departments, diagnostic testing facilities, sequencing laboratories, and organizations that can analyze large genomic data sets, leading to complex operational considerations.^13, 14^ Staffing shortages in diagnostic laboratories can be exacerbated by increases in community transmission of SARS-CoV-2 due to additional diagnostic workloads and infections requiring isolation among staff, leading to delays in specimen submission and impacting the timeliness of results. Additionally, the surveillance program must consider how sequencing results will be interpreted by the public. Misinformation has impeded SARS-CoV-2 pandemic responses and genomic surveillance findings may be misconstrued by the public and the media.^7, 15^ Thus, coordination among genomic surveillance programs and public health departments for interpretation and effective communications leveraging trusted sources are also critical.

The emergence of concerning variants often requires urgent action by genomic surveillance programs to aid public health response. Here we describe a response playbook which includes guidance for adjusting surveillance strategies, laboratory processes, public health communications and engagement based on known characteristics of the variant and local community risk. Additionally, we describe the application of the playbook response to surveillance of Omicron BA.1 and compare the correlation and timeliness of WGS and genotyping PCR.

## Methods

### Study Setting and Participants

The Regional Innovative Public Health Laboratory (RIPHL) is a public-academic partnership between the Chicago Department of Public Health (CDPH) and Rush University Medical Center (RUMC). RIPHL was established to provide advanced molecular surveillance capacity to CDPH, which does not operate its own public health laboratory. RIPHL accepts remnant clinical SARS-CoV-2-positive nasopharyngeal or anterior nasal swab specimens from 14 hospital and health care systems in Chicago, referred to hereafter as”partners”. At baseline, partners are encouraged to submit a convenience sample of 15 remnant specimens per week. Specimens are grouped into a single batch each week for processing. With ∼12 partners submitting 15 specimens weekly and an expected 80% yielding WGS, the surveillance system aims to generate around 150 sequences per week — sufficient to provide 95% confidence of detecting a new lineage within one week of its causing 2% of incident cases.^16^ At times, specimens from individuals with specific epidemiologic features are requested. This focused surveillance aims to enrich datasets with characteristics of interest to identify signals of increased virulence, immune evasion, or transmissibility — for example, cases of SARS-CoV-2 among fully vaccinated individuals, people with severe COVID-19 symptoms or disease outcome including death, and individuals living or working in a skilled nursing facility.

### Specimen processing

Upon receipt at RIPHL, specimens are heat inactivated then undergo nucleic acid extraction using a Maxwell RSC (Promega). RT-qPCR is performed using UltraPlex 1-Step ToughMix (Quantabio) and N2 primer/probe mixes (CDC 2019-Novel Coronavirus Real-Time RT-PCR Diagnostic Panel, IDT) are used to quantify SARS-CoV-2 RNA. Extracted samples undergo reverse transcription using High Capacity Reverse Transcription Kit (Applied Biosystems). Library preparation is performed using the XGen SC2 Expanded Amplicon Panel kit (Integrated DNA Technologies). Pooled libraries are sequenced on an Illumina NovaSeq6000 at an external sequencing core facility. For small-scale rapid internal sequencing, pooled libraries are sequenced on Illumina iSeq or MiniSeq.

### Bioinformatics

Sequencing data is analyzed using the TheiaCoV pipeline (Theiagen Genomics).). Briefly, raw read quality is assessed and host reads are removed. Reads are quality-and adapter-trimmed and aligned to reference genome (MN908947.3). Primer trimming, consensus assembly, and variant calling is performed using iVar^17^. SARS-CoV-2 lineage designation and quality assessment is performed using pangolin v4.0.6-pdata-1.8 and NextClade1^18,19^.

### RT-qPCR genotyping (“variant PCR”)

TaqMan SARS-CoV-2 Mutation Panel Assays (ThermoFisher) were used according to manufacturer’s instructions in 384-well format using 5μl RNA extract. For Omicron BA.1, K417N (present in Omicron but not present in Delta) was selected and L452R (present in Delta but not in Omicron) was included to confirm the genotype, as a small percentage (∼0.2%) of Delta strains contained the K417N mutation. Specimens were designated Omicron if they were K417N positive and L452R negative and designated Delta if they were L452R positive and K417N negative. Specimens positive for both K417N and L452R were designated as Delta +K417N. These assays were selected as they appeared to be defining mutations of each lineage, present in a large proportion of available sequence data for each lineage, and due to their availability as stocked commercial assays for rapid procural. Other commercial genotyping assays targeting regions with several mutations in Omicron were not selected, as these neighboring mutations could potentially affect assay efficiency.

### Variant Response Playbook

The RIPHL Variant Response Playbook (“Playbook”) is structured into three progressive tiers, titrated according to assessments of the need for enhanced genomic surveillance. Each of the Playbook’s tiers include actions across the following domains: i) sampling strategy, ii) laboratory processing, iii) public health department coordination, and iv) partner engagement (Table 1).

**Table 1:** Regional Innovative Public Health Laboratory’s Variant Response Playbook by Tiers and Key Domains.

| **Variant Designation** | **Tier 1** | **Tier 2** | **Tier 3** |
| --- | --- | --- | --- |
| Characteristic | Numerous or concerning mutations that may affect viral fitness | Mutations that are likely to cause biological changes or increased fitness | Mutations cause concerning biological changes and significantly increased fitness |
| Growth | Increasing in prevalence in one region | Increasing in prevalence in multiple global locations | Increasing in prevalence in nearby locations |
| Designation | Variant of interest, variant under investigation, variant being monitored | Variant of concern | Variant of concern, variant of high consequence |
| Public health consequence | Minor consequence suspected | Moderate consequence suspected | Significant consequence suspected |
| Example | BA.2.75 | BA.5 | BA.1 |
| **Playbook Response** | **Tier 1** | **Tier 2** | **Tier 3\*** |
| Sampling strategy | Maintain baseline sampling strategy | Prioritize surveillance samples by most recent collection date and/or epidemiologic links to new variant | Increased weekly sample submissions |
| Laboratory processing | Purchase variant PCR reagents | Implement variant PCR screening | Implement rapid WGS of VOCs |
| Public health coordination | Communicate variant properties, emerging research, and transmission patterns | Integrate internal results with other data sources; begin public-facing communications | Expedite epidemiologic analysis of vaccine breakthrough, reinfection, hospitalization, death |
| Partner engagement | Maintain baseline quarterly and upon request reporting | Communicate implementation of Playbook; prepare for increased weekly sample submissions | Send weekly reports; Communicate increase in weekly sample submissions |

\*Tier 3 includes Tier 2 activities

#### Tier 1

Playbook Tier 1 is implemented when a new variant is reported anywhere in the world which a) contains numerous or concerning mutations that are predicted to affect viral fitness, such as transmissibility, or immune evasion; b) is significantly increasing in prevalence in one international region; or c) is classified as a variant of interest, variant under investigation, or variant being monitored by national or global health agencies.

i. Sampling strategy. Baseline sampling strategy is maintained.
ii. Laboratory processing. Variant-specific qPCR reagents are identified are procured, or designed if existing reagents are not available. As custom qPCR probes have lengthy delivery times and require validation, commercially available assays are preferred. Supplementary laboratory staffing for qPCR screening is arranged.
iii. Public health department coordination. Variant summaries, including variant genome properties, growth rates where the variant has been observed, and emerging research on the variant and/or associated mutations, are distributed by subject matter experts within the academic-public health working group. Public-facing communications (e.g., responding to media and public queries) are prepared.
iv. Partner engagement. Summary reports about variant incidence and prevalence are provided to submitting partners quarterly.

#### Tier 2

Playbook Tier 2 is implemented when a new variant a) contains suspected changes to its biological properties (e.g., significant immune escape, decreased effectiveness of therapeutics, increased transmissibility), b) is rapidly increasing in prevalence in multiple global locations, or is classified as a variant of concern by national or global health agencies. Tier 2 focuses on the rapid identification of variant introduction and/or rapidly changing variant proportions and enacts processes that were planned in Tier 1.

i. Sampling strategy. Specimen batching protocols are adjusted to prioritize routine surveillance samples by most recent collection date. Older samples (specimen collection date >14 days prior) are stored for future processing if weekly laboratory capacity is reached. In addition, targeted cases with epidemiologic links to the new variant are prioritized. Specimens undergoing expedited analysis are tagged as”targeted surveillance” and not”baseline surveillance” when submitting to public repositories to avoid skewing variant estimates.
ii. Laboratory processing. Variant PCR is performed on all surveillance specimens. In-house sequencing of new variants identified by variant PCR is performed to confirm lineage until the accuracy of PCR is confirmed. Increased laboratory staffing to perform additional qPCR and analyze data is implemented to prevent delays to WGS. Supplementary and alternative redundant staffing is arranged to address throughput, inclusion of the qPCR assay, and staff absenteeism due to a potential COVID-19 surge.
iii. Public health department coordination. Other sources of variant proportion information, e.g. CDC National SARS-CoV-2 Surveillance System^20^ and state or regional GISAID submissions^21^, are accessed and analyzed weekly in comparison to internal data. Executive public health leadership is briefed, and proactive public-facing communications are developed.
iv. Partner engagement. Communication to partnering laboratories and health organizations include descriptions of the new variant and playbook implementation. Presentations to local hospital epidemiologists, clinicians, and microbiologists are given upon request to educate local healthcare experts on the new variant, public health significance, and local emergence.

#### Tier 3

Tier 3 begins when a variant has concerning properties compared to existing variants which may have significant public health consequences or require public health policy changes (e.g., poor clinical response to existing therapies or vaccines, increased disease severity or outcomes, expansion of susceptible populations, or high levels of community transmission). The variant may be designated a variant of high consequence (VOHC) and detected in a geographically proximate location, e.g. detected in the United States. Tier 3 represents intensified surveillance for increased sensitivity of variant detection and proportion monitoring for variants with higher potential public health consequence compared to Tier 2 variants. All processes activated in Tier 2 continue.

i. Sampling strategy. The number of clinical specimens analyzed by WGS and variant PCR is doubled to ∼300 to provide 95% confidence of detecting a new lineage causing 1% of incident cases.^15^
ii. Laboratory processing. Workflow and staffing adjustments for increased specimen submissions for both variant PCR and WGS are implemented. Specimens are processed in daily batches (as opposed to weekly) to prevent delay in variant PCR or WGS detection, i.e. samples extracted on Day 1 undergo variant PCR and WGS library preparation on Day 2, specimens extracted on Day 2 undergo variant PCR and WGS library preparation on Day 3, etc. In-house sequencing on small scale sequencers (e.g., Illumina iSeq, MiniSeq, or MiSeq) is performed same-day for all new variant specimens identified by variant PCR for early investigation of lineages, cluster detection, and support of transmission links, until sequencing capacity on small in-house sequencers is exceeded.
iii. Public health department coordination. Health department expedites linkage of vaccination, hospitalization, epidemiologic, and prior infection data across multiple information systems to WGS results and determines variant-associated risks. Decision makers outside of the public health department including elected officials are briefed. Communications of these results are developed for local government officials, health care professionals and the public, as appropriate.
iv. Partner engagement. Partners increase weekly specimen submissions. Where feasible, partners submit SARS-CoV-2 specimens daily rather than weekly. Weekly variant PCR and WGS reports are sent to partners. Additionally, internal variant data and relevant external regional data are presented to partners, local hospital epidemiologists, and public health department leadership weekly.

#### Step-down

Step-down procedures are implemented when enhanced surveillance is no longer needed for more rapid or granular surveillance, e.g. when sufficient data is collected to track variant emergence and estimate variant growth rate and associated health outcomes, or if the variant fails to outcompete other circulating strains or does not emerge locally. Upon step-down, variant PCR is no longer performed. Partners are instructed to reduce the number of samples submitted to baseline levels. Data analysis and reporting returns to baseline.

## Results

### Playbook Implementation for Enhanced Genomic Surveillance

On November 26, 2021, when the WHO classified Omicron as a Variant of Concern (VOC), RIPHL initiated the first steps of the Playbook’s Tier 1 (Figure 1). Omicron sequences were compared to currently circulating strains, unique mutations were identified, and commercially available genotyping kits to Omicron-specific mutations were ordered. On November 30, 2021, as Omicron BA.1 was rapidly increasing in prevalence around the globe, suggesting significant transmission advantage over previous variants, RIPHL entered Tier 3. CDPH issued a Health Alert to request RIPHL partners to double their submissions on November 30, 2021,^22^ and submissions increased from an average 127 specimens per collection week in October and November 2021 (MMWR 39 – 47) to 261 per week from December 2021 to February 2022 (MMWR 48 – 5) (Figure 2). Tier step-down was enacted on Jan 27, 2022, as data indicated local Omicron BA.1 prevalence exceeded 90% and enhanced surveillance was no longer needed (Figure 1).

**Figure 1.**
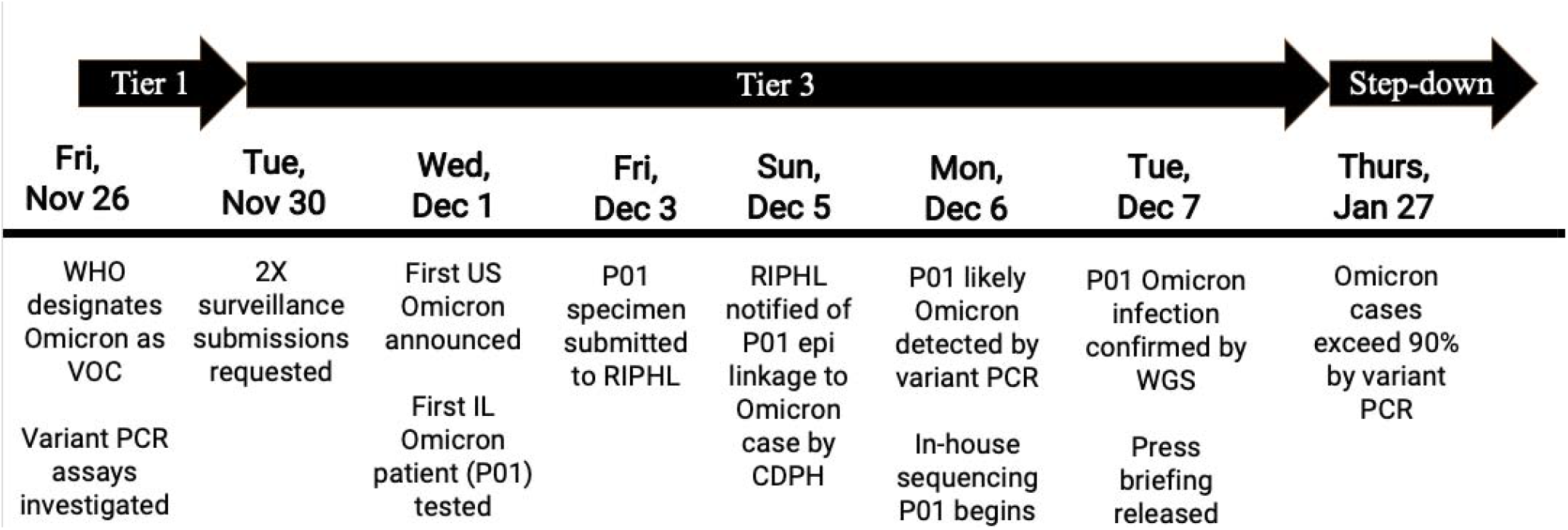
Timeline of Omicron BA.1 Playbook response. Timeline and summary of playbook implementation in response to Omicron BA.1 and detection of the first Omicron case in Illinoi are summarized by date. Key events surrounding tier enactment and the detection of the first Omicron case in Illinois are highlighted.

**Figure 2.**
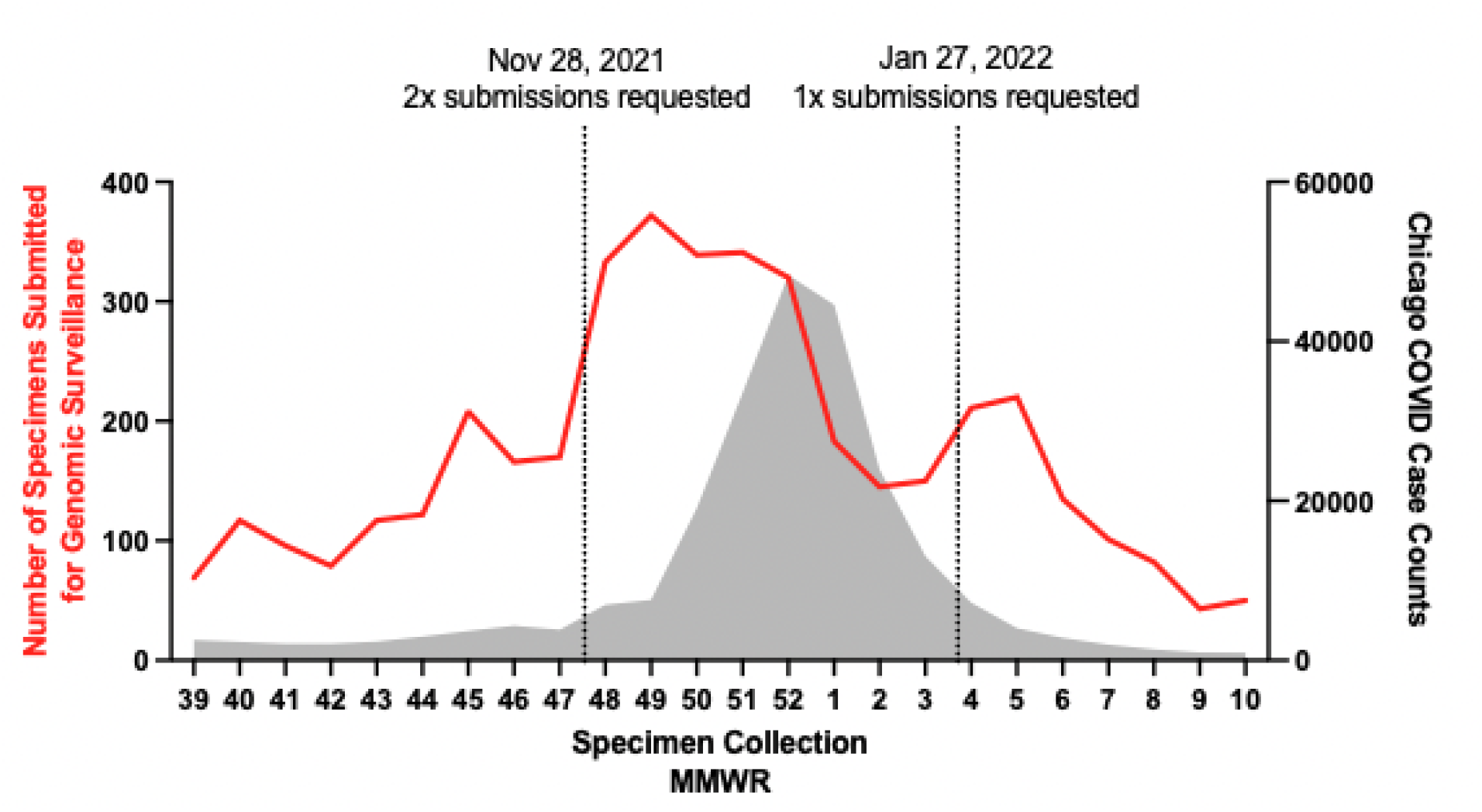
Samples Received During Omicron BA.1 Wave and Tier 3 Response, October 2021 – March 2022. The number of SARS-CoV-2 specimens submitted for genomic surveillance is shown (red line, left axis), aggregated by specimen collection week. Overlaid are Chicago COVID-19 case counts (grey, right axis). The date when Playbook changes to specimen submission numbers were requested are shown by vertical lines. The decline in specimen around 2022 MMWR 1-3 was attributed to stress on clinical laboratories during the period of high cases, which limited their ability to submit samples to RIPHL. Data source: Chicago Department of Public Health, Regional Innovative Public Health Laboratory.

### Rapid Detection of the First Omicron Case by Variant PCR and WGS

Playbook responses enabled rapid identification of the first Omicron case in Illinois (Figure 1).^23^ First, doubling specimen numbers increased the probability that the specimen was submitted for genomic surveillance. Second, daily rather than weekly submission of specimens by select partners, daily batching of specimens, and prioritization of cases with epidemiologic linkages to Omicron cases and more recently collected specimens ensured timely processing of this specimen. Third, variant PCR provided evidence of a likely Omicron genotype only one business day after submission. Fourth, in-house sequencing provided WGS confirmation of the Omicron genotype within 24 hours of identification via variant PCR. Fifth, increased communications between the laboratory and health department allowed for specimens with epidemiologic linkages identified through contact tracing to be prioritized.

### Variant PCR Screening

Variant PCR was performed on all specimens collected between November 28, 2021 and January 27, 2022. 1,657 of 1,833 samples (90.4%) successfully underwent variant PCR and received a lineage determination, while fewer (1,560 of 1,833; 85.1%) were successfully sequenced. To determine the accuracy of variant PCR, the results were compared to the definitive WGS lineage result (Table 2). Both variant PCR) and WGS lineage results were available for 1,541 samples, and concordant results were reported in nearly all cases (1,539; 99.9%) (Table 3, Cohen’s kappa = 0.997, 95% CI 0.994 - 1.000). The two discordant samples were both called Delta by variant PCR and Omicron by WGS. Of these, one showed both Delta and Omicron genomic regions by WGS, with Delta genotypes at the K417N and L452R mutation sites. Further investigation revealed tjat this specimen was likely an Omicron-Delta co-infection. The other discordant sample could not be explained upon investigation of sequence data. Overall, variant proportions determined by variant PCR were nearly indistinguishable from WGS (Figure 3).

**Table 2.**
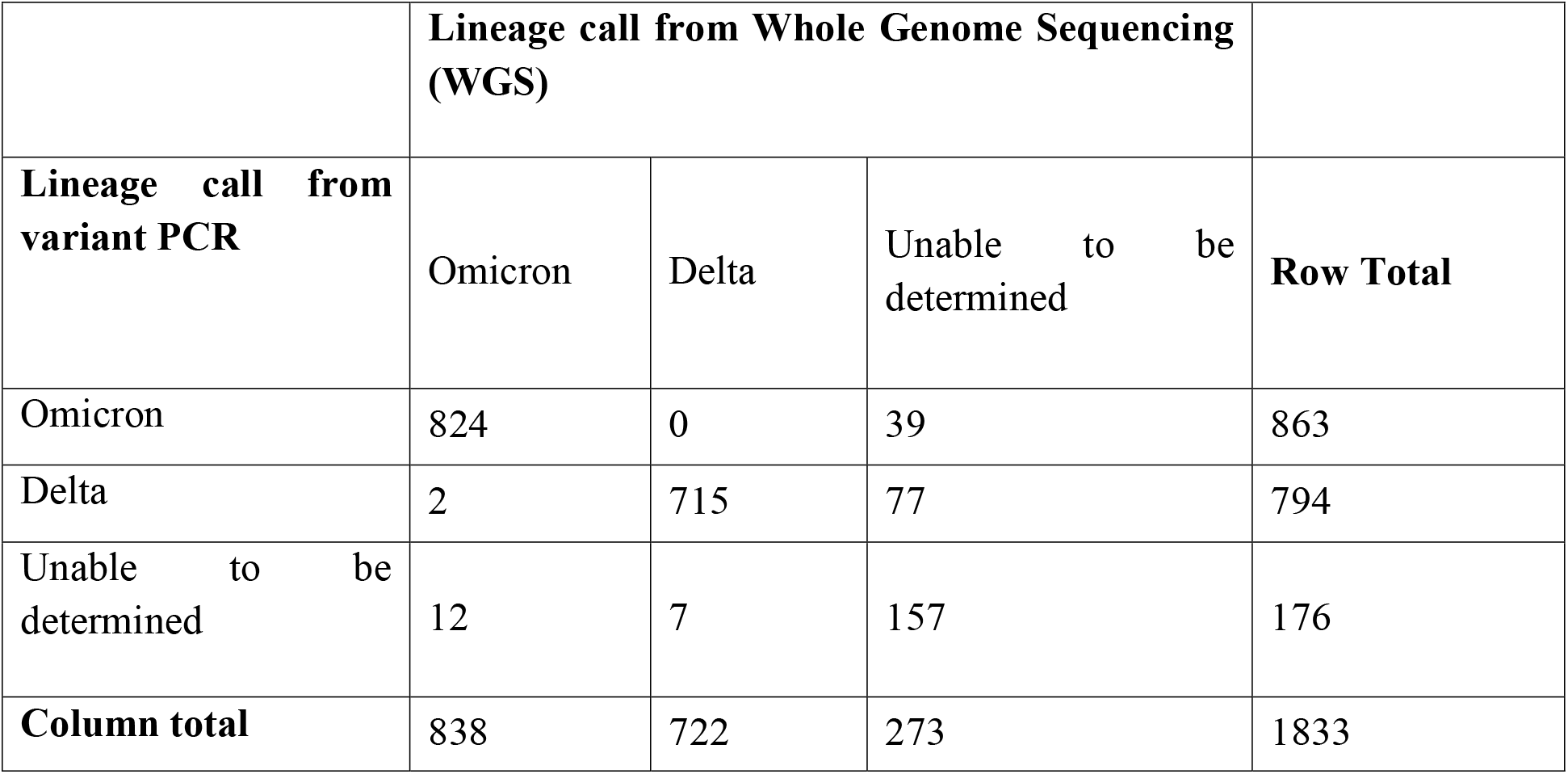
Comparison of variant PCR and WGS lineage results for samples received December 2, 2021 - January 21, 2022.

|  | **Lineage call from Whole Genome Sequencing (WGS)** |  |  |  |
| --- | --- | --- | --- | --- |
| **Lineage call from variant PCR** | Omicron | Delta | Unable to be determined | **Row Total** |
| Omicron | 824 | 0 | 39 | 863 |
| Delta | 2 | 715 | 77 | 794 |
| Unable to be determined | 12 | 7 | 157 | 176 |
| **Column total** | 838 | 722 | 273 | 1833 |

**Figure 3.**
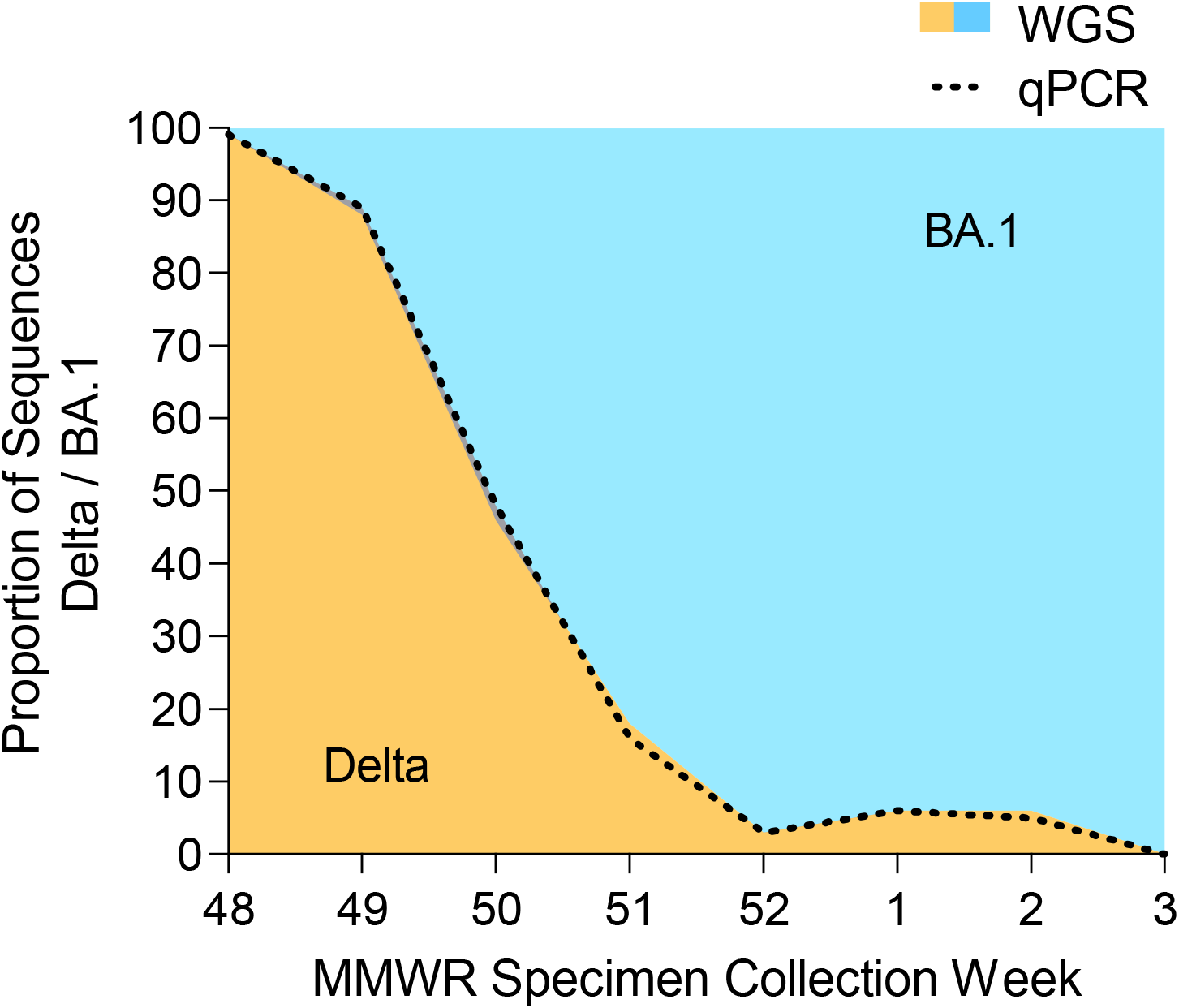
Comparison of Delta/BA.1 lineage proportions by WGS and variant PCR. The proportion of sequences determined to be Delta (orange) or Omicron BA.1 (blue) by WGS is plotted, with the proportion of sequences determined to be Delta by variant PCR shown by dotted line; specimens collected November 28, 2021 – January 21, 2022.

Variant PCR was more sensitive than WGS, reducing the number of undetermined lineage calls from 273 to 176, a 35.5% reduction. The median time from specimen submission to RIPHL to lineage call was 7 days for variant PCR and 27 days for WGS, a reduction of 71.4%. The long WGS turnaround times were in part due to this timeframe spanning the winter holiday, during which staffing shortages delayed library preparation and the sequencing facility was closed for one week. For samples processed before the holiday closure (N=274), variant PCR results were available a median 6 days after specimen arrival at RIPHL, compared to 19 days for WGS.

## Discussion

Using a tiered Playbook to address emerging SARS-CoV-2 variants allowed for a structured but agile approach to changes in genomic surveillance. Having identifiable triggers to move from baseline to enhanced workflows and stepdown allowed for clear internal communication and expectations from the laboratory, clinical partners, and the public health department. Further, communications of actionable data to the Chicago healthcare community 1) increased local understanding of genomic surveillance, 2) recruited additional hospitals submitting to RIPHL, increasing its demographic coverage, and 3) helped inform which data is helpful when making public health responses to emerging variants.

The introduction of variant PCR was critically important to the success of the Playbook response to BA.1. Variant PCR was highly accurate, faster, and more sensitive than WGS, providing earlier estimates of variant proportions and additional genotype data for specimens that were unable to be sequenced by WGS. Variant PCR also proved easier to scale-up during Tier 3 and was less susceptible to staffing shortages and core facility delays, due to qPCR requiring less highly trained staff, less preparation and equipment run time, and being run in-house without external facility lead time. Variant PCR proved highly useful to perform timely local variant fitness estimates for the new variant, such as growth rate and advantage.^24^ One limitation of variant PCR is the availability of off-the-shelf assays targeting unique mutations on future variants. While custom assays can be developed, the time for production and validation might preclude its usefulness for rapid detection. To overcome this, genomic surveillance programs might consider stocking assays targeting several mutations, or a multiplex panel capable of distinguishing diverse variants.^25^ Although, genotyping qPCR assays only need to differentiate the growing lineage relative to the contemporaneous lineages to be effective, therefore, off-the-shelf assays are frequently available for this purpose.

A limiting factor for increased surveillance was the capacity of submitting laboratories during a COVID-19 surge to send additional specimens as they are facing staffing shortages and an increased diagnostic testing demand. Indeed, during the first three weeks of 2022, we observed a seemingly paradoxical decline of specimen submissions with increasing SARS-CoV-2 cases (Figure 2). Upon consultation with submitting laboratories, the declined specimen submission was due to stress on lab personnel who were performing a high volume of diagnostic testing while also experiencing staffing shortages due to COVID-19 infections. To address the challenge in the future, opportunities for the genomic surveillance program and the health department to provide support, including staffing resources to collect, process, and transport specimens is being investigated. Daily submission of specimens requested in Tier 3 was only feasible for a clinical laboratory located at the same institution as RIPHL, underscoring the value of co-locating a molecular surveillance lab with a large clinical provider.

While the playbook described here adapts the specific baseline genomic surveillance processes of RIPHL, it could also serve as a template for other genomic surveillance programs to adapt and escalate their baseline laboratory, sampling, and operations processes to quickly respond to new SARS-CoV-2 variants or variants of other relevant pathogens. Additionally, the playbook could be expanded to include more specific action items, such as specific analyses that are run at defined frequencies, or internal and public report templates, to further ease the coordination needed to respond to new variants swiftly and comprehensively.

## Conclusions

A strong regional genomic surveillance system can enable local public health departments to form swift and data-driven responses to emerging pathogen variants. Regional genomic surveillance programs can benefit from defining prescribed responses to new variants, as these variants are often accompanied by increases in testing, data requests, and epidemiologic analyses which can burden already complex processes. Different levels of response are warranted by different variants, depending on the predicted variant properties, clinical consequences (e.g., hospital and emergency department burden), and public health response that might be required. For example, the Omicron BA.1 surge triggered indoor masking mandates, large gathering restrictions, travel guidelines, and monoclonal antibody therapy recommendations in many jurisdictions. In contrast, Omicron BA.4 and BA.5 received less substantial public health responses. Here our 3-tiered playbook represents an outline of progressive strategies for specimen sampling, laboratory processing, communication with specimen submitters, and coordination with the local public health department depending on new variant properties.

Key improvements to standard genomic surveillance were observed in response to Omicron BA.1. First, genomic surveillance programs can benefit from using variant qPCR genotyping as a complement to WGS for accurate, sensitive, and rapid monitoring of novel variant proportions. Second, increasing the number of submitted specimens can enable more sensitive and rapid detection of emerging variants. Third, increased communication and coordination between the laboratory and public health department can ensure prompt reporting to public health leadership and informing the public.

## Data Availability

The WGS datasets generated and/or analysed during the current study are available in GISAID, http://gisaid.org, under accession EPI_SET_230901ya. The qPCR datasets used and/or analysed during the current study are available from the corresponding author on reasonable request.

## List of Abbreviations

SARS-CoV-2: Severe acute respiratory syndrome coronavirus 2
WGS: Whole genome sequencing
PCR: Polymerase chain reaction
COVID-19: Coronavirus disease 2019
VOC: Variant of concern
RIPHL: Regional Innovative Public Health Laboratory
CDPH: Chicago Department of Public Health
RUMC: Rush University Medical Center
RT: Reverse Transcription
GISAID: Global Initiative on Sharing All Influenza Data
VOHC: Variant of High Concern
WHO: World Health Organization
MMWR: Morbidity and Mortality Weekly Report week
CI: Confidence interval

## Declarations

### Ethics approval

The project was determined to not meet the definition of human subjects research and was exempt from ethical approval by the Rush University Institutional Review Board.

### Consent for publication

Not applicable

### Authors’ contributions

Authors HJB, AK, IG, and MKH conceived the study. Authors HB, RT, XQ and SJG developed methods. Authors HJB, SB, and KE prepared data and performed analyses. Authors RT, IG, MP, and MKH supervised the study. Authors HJB, AK, and IG wrote the original draft. All authors read and approved the final manuscript.

## Acknowledgements

The authors would like to thank the Regional Innovative Public Health Laboratory partner organizations: Advocate Illinois Masonic Medical Center, Advocate Trinity Hospital, Ascension, Ann & Robert H. Lurie Children’s Hospital of Chicago, John H. Stroger, Jr. Hospital, Loyola University Medical Center, Northwestern Memorial Hospital, Rush University Medical Center, Sinai Health System, UChicago Medicine, University of Illinois Chicago.

